# The Genomic Landscape of Rare Disorders in the Middle East

**DOI:** 10.1101/2022.09.17.22279590

**Authors:** Maha El Naofal, Sathishkumar Ramaswamy, Ali Alsarhan, Ahmed Nugud, Alan Taylor, Ruchi Jain, Nour Halabi, Sawsan Yaslam, Roudha Alfalasi, Shruti Shenbagam, Martin Bitzan, Lemis Yavuz, Deena Wafadari, Hamda Abulhoul, Shiva Shankar, Munira Al Maazmi, Ruba Rizk, Zeinab Alloub, Haitham Elbashir, Mohamed O. E. Babiker, Nidheesh Chencheri, Ammar AlBanna, Meshal Sultan, Mohamed El Bitar, Safeena Kherani, Nandu Thalange, Sattar Alshryda, Roberto Di Donato, Christos Tzivinikos, Ibrar Majid, Alexandra F. Freeman, Corina Gonzalez, Arif O. Khan, Hisham Hamdan, Walid Abuhammour, Mohamed Al Awadhi, Abdulla Al Khayat, Alawi Alsheikh-Ali, Ahmad N. Abou Tayoun

## Abstract

**BACKGROUND:** Rare diseases collectively impose significant burden on healthcare systems, especially in underserved regions, like the Middle East, which lack access to genomic diagnostic services and the associated personalized management plans.

**METHODS:** We established a clinical genomics and genetic counselling facility, within a multidisciplinary tertiary paediatric center, in the United Arab Emirates to locally diagnose and manage patients with rare diseases. Clinical genomic investigations included exome-based sequencing, chromosomal microarrays, and/or targeted testing. We assessed the diagnostic yield and implications for clinical management among this population.

**RESULTS:** We present data on 529 patients with rare diseases (47% females; Average age, 4 years) representing 41 countries primarily from the Arabian Peninsula, the Levant, Africa, and Asia. The cumulative diagnostic yield was 35.7% (95% CI, 31.8% - 39.9%) and was higher for genomic sequencing-based testing than chromosomal microarrays (41.2% versus 16.9%, P=0.0001) across all indications, consistent with the higher burden of single gene disorders. Of the 124 Mendelian disorders identified in this cohort, the majority (N = 110) were encountered only once, and those with recessive inheritance accounted for ~60% of sequencing diagnoses. Of patients with positive genetic findings (N = 189), 71.9% were less than 5 years of age, and 65.6% were offered modified management and/or intervention plans. Interestingly, 18.5% of patients with positive genetic findings received delayed diagnoses (age range 7 – 37 years), most likely due to lack of access to genomic investigations in this region. One such genetic finding ended over a 10-year long diagnostic odyssey, leading to a life-threatening diagnosis in one patient, who was then successfully treated using an experimental allogenic bone marrow transplant. Finally, we present cases with candidate genes within regions of homozygosity, likely underlying novel recessive disorders.

**CONCLUSIONS:** Early access to genomic diagnostics for patients with suspected rare disorders in the Middle East is likely to improve clinical outcomes while driving gene discovery in this historically underrepresented population.

## Introduction

Although individually rare, the cumulative prevalence of the roughly 7000 known rare diseases can be as high as 6%; nearly 450 million individuals may be affected globally^1,2^. Despite the fact that most of these diseases have genetic origins, affected patients go through extended diagnostic odysseys averaging 6 – 8 years, characterized by multiple hospitalizations, unnecessary diagnostic workup, and/or inefficient management or treatment plans, leading to substantial social and economic burden on families and healthcare systems^3,4^.

The prevalent close relative marriages and large family structures in populations of the Middle East are expected to lead to high rates of rare Mendelian disorders^5^. On the other hand, the lack of specialized care centers and the limited access to genomic services^6^ are likely to contribute to longer diagnostic odysseys than have been described elsewhere, resulting in missed opportunities for personalized management plans.

To address this gap, we established a dedicated clinical genomics center in one of the first standalone children’s specialty hospitals in the Middle East, Al Jalila Children’s Specialty Hospital (AJCH). Unlike previous studies which focused on specific homogeneous populations (mainly Saudis^7^ and Qataris^8^) from this region, our center is located within Dubai, a regionally accessible city with high population diversity, which enabled us to recruit, genetically diagnose, and care for patients with rare diseases from underserved populations, representing at least 41 countries of the Middle East, Africa, and Asia, which have historically been underrepresented in genetic studies.

## Methods

### Study Design and Participants

This study includes patients referred for clinical genomic testing from April 2019 - December 2021. Patients’ physicians ordered the tests, and the physician or a certified genetic counsellor explained the benefits, limitations, and risks of testing, obtaining written informed consent.

Peripheral blood samples were obtained from each patient and, in the case of trio whole exome sequencing, their parents. Clinical data were provided by the referring physician either through the electronic medical record (internal patients) or on the requisition forms (outside patients). All clinical, demographic, and genetic data were summarized and reported by the laboratory molecular geneticist. All patient samples were de-identified for this report. Patient IDs in supplementary tables were newly assigned and are only known to the research group. This study was reviewed by Dubai Healthcare Authority Research Ethics Committee (AJCH – 44) and determined to meet exemption criteria with waiver of informed consent. Patient #297 was consented to share additional details and to publish this work. This study followed the Strengthening the Reporting of Observational Studies in Epidemiology (STROBE) reporting guideline^9^.

### Genomic Investigations, Bioinformatic Analysis, and Sequence Variant Interpretation and Reporting

All testing was performed in a College of American Pathologists (CAP)-accredited genomics facility. Sample preparation and analyses were performed as previously described^10^ and summarized in **Supplementary Methods**^11–13^ (available upon request).

All retained sequence variants were classified following the American College of Medical Genetics and Genomics (ACMGG) sequence variant interpretation guideline^14^. Pathogenic and likely pathogenic variants in genes relevant to the patients’ primary indications were reported and were considered diagnostic if patient’s phenotype (based on physician’s notes and feedback), disease mechanism, and inheritance were all consistent. Variants of uncertain significance relevant to patients’ primary indications were also reported, though were not considered diagnostic.

When identified, medically actionable secondary findings were reported if patients opted in for such findings. Only pathogenic or likely pathogenic variants in the 59 genes recommended by ACMGG^15^ were reported.

### Clinical Indications

The indications for genomic testing were categorized into 11 major groups based on involved systems(s). Since many patients have a combination of neurological and neurodevelopmental disorders, we combined both into a single group. For ‘complex multiple systemic involvement’, we gathered all the cases that can be under more than 2 groups. Finally, for a minority of patients who did not fit in any category, we grouped them as “others”.

### Clinical Utility

For patients with diagnostic findings, we documented the changes in the management plans – either management or intervention – that occurred or were recommended after obtaining the genetic results. ‘Intervention’ is defined as any planned process applied to the patient that requires medication introduction, change or discontinuation, diagnostic imaging, further testing, or surgical procedure. ‘Management’ is defined as any planned process applied to the patient that does not fit the definition of “intervention,” including recommendations or advice, referral to other services, or follow up plans.

### Statistical Analyses

Variables, mainly diagnostic yields of different testing modalities, were compared using the Fisher exact test. Tests were 2-tailed, and P < .05 was considered statistically significant.

## Results

### Demographic and Clinical Characteristics of Patients

Five hundred and twenty-nine patients (47% females; average age, 4 years) representing 41 countries (**Supplementary Table 1**) in the Arabian Peninsula (58.4%), the Levant (11.5%), Africa (9.6%), Asia (12.5%), and other geographic regions (8.1%) (**Figure 1A-B** and **Table 1**) either presented (N = 425) or had their genetic testing referred (N = 104) to AJCH. Most patients (48.2%) were between 0 – 2 years, while 19.3% were 2 – 5 years and 32.5% ≥5 years of age (**Table 1**).

**Table 1.** Patient Demographic Information and Ancestry

| Arabs |  |  |  |  |  |  |  |  |  | Non-Arabs |  |  |  |  |  |
| --- | --- | --- | --- | --- | --- | --- | --- | --- | --- | --- | --- | --- | --- | --- | --- |
|  | All<br>(n; %) | Arabian Peninsula |  | Levant |  | North Africans |  | Others Unspecified |  | Asians |  | Europeans/North Americans |  | Africans/Others |  |
|  |  | Total no. | Proportion%<br>(95% CI) | Total no. | Proportion%<br>(95% CI) | Total no. | Proportion%<br>(95% CI) | Total no. | Proportion%<br>(95% CI) | Total no. | Proportion%<br>(95% CI) | Total no. | Proportion%<br>(95% CI) | Total no. | Proportion%<br>(95% CI) |
| Age, y |  |  |  |  |  |  |  |  |  |  |  |  |  |  |  |
| 0 - 2 | 255; 48.2% | 140 | 54.9%<br>(48.8 - 60.9) | 29 | 11.4%<br>(8.0 - 15.9) | 18 | 7.1%<br>(4.5 - 10.9) | 14 | 5.5%<br>(3.3 - 9.0) | 37 | 14.5%<br>(10.7 - 19.4) | 10 | 3.9%<br>(2.1 - 7.1) | 7 | 2.7%<br>(1.3 - 5.6) |
| 2 - 5 | 102; 19.3% | 61 | 59.8%<br>(50.1 - 68.8) | 14 | 13.7%<br>(8.4 - 21.7) | 7 | 6.9%<br>(3.4 - 13.5) | 6 | 5.9%<br>(2.7 - 12.2) | 8 | 7.8%<br>(4.0 - 14.7) | 4 | 3.9%<br>(1.5 - 9.7) | 2 | 2.0%<br>(0.5 - 6.9) |
| ≥5 | 172; 32.5% | 108 | 62.8%<br>(55.4 - 69.7) | 18 | 10.5%<br>(6.7 - 15.9) | 16 | 9.3%<br>(5.8 - 14.6) | 4 | 2.3%<br>(0.9 - 5.8) | 21 | 12.2%<br>(8.1 - 17.9) | 5 | 2.9%<br>(1.2 - 6.6) | 0 | 0 |
| Sex |  |  |  |  |  |  |  |  |  |  |  |  |  |  |  |
| Male | 280; 52.9% | 173 | 61.78%<br>(56.0 - 67.3) | 31 | 11.1%<br>(7.9 - 15.3) | 23 | 8.2%<br>(5.5 - 12.0) | 12 | 4.3%<br>(2.5 - 7.3) | 28 | 10.0%<br>(7.0 - 14.1) | 9 | 3.2%<br>(1.7 - 6.0) | 4 | 1.4%<br>(0.6 - 3.6) |
| Female | 249; 47.1% | 136 | 54.6%<br>(48.4 - 60.7) | 30 | 12.0%<br>(8.6 - 16.7) | 18 | 7.2%<br>(4.6 - 11.1) | 12 | 4.8%<br>(2.8 - 8.2) | 38 | 15.3%<br>(11.3 - 20.3) | 10 | 4.0%<br>(2.2 - 7.2) | 5 | 2.0%<br>(0.9 - 4.6) |
| Overall | 529 | 309 | 58.4%<br>(54.2 - 62.5) | 61 | 11.5%<br>(9.1 - 14.5) | 42 | 7.8%<br>(5.8 - 10.3) | 24 | 4.5%<br>(3.1 - 6.7) | 66 | 12.5%<br>(9.9 - 15.6) | 20 | 3.6%<br>(2.3 - 5.5) | 9 | 1.7%<br>(0.9 - 3.2) |

**Figure 1.**
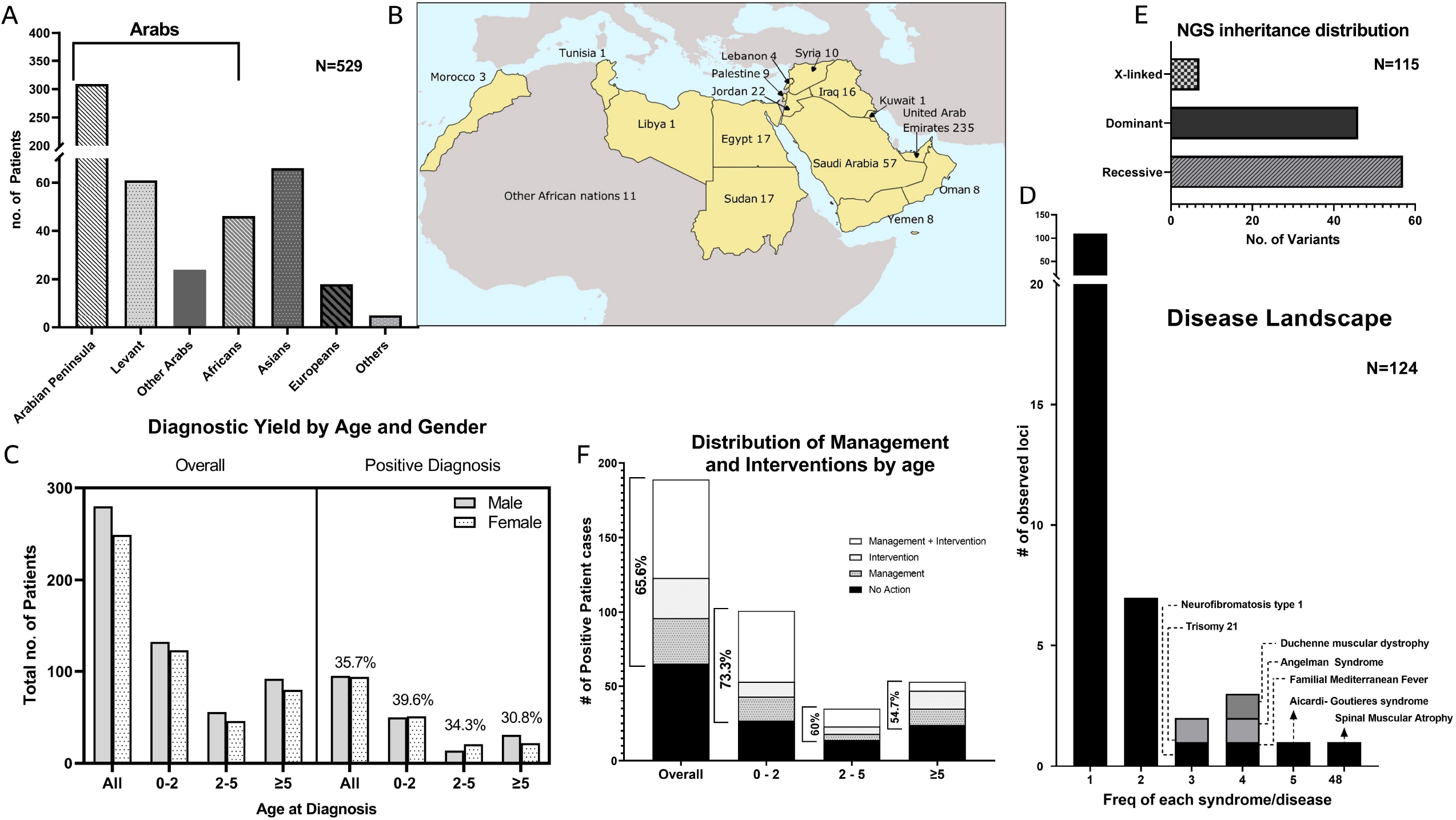
Cohort Summary. A) Distribution of patients’ origins by geographical regions. The bracket denotes patients of Arab origins, in the Middle East and North Africa, whose distribution by country is shown in (B). Note that other Arabs (N = 20) of unknown countries of origin in (A) are not included on the map. C) Distribution of patients and diagnostic yield by age and gender. D) Frequencies of rare diseases (N = 124) observed in this cohort. Most diseases (>100) are seen only once. E) Breakdown of inheritance patterns of cases with positive findings by sequencing. F) Age distribution of patients with positive findings who received modified management and/or intervention plans.

Overall, 60.5% of patients presented with neurological phenotypes (**Table 2**). Comprehensive genomic sequencing-based testing (50.5%), in the form of whole or indication-based exome sequencing (**Supplementary Table 2**), or chromosomal microarrays (30%) were the most common clinical genomic investigations. Targeted testing, mainly for spinal muscular atrophy (SMA), fragile X, or methylation disorders, was performed for 37% of patients. Around 16% of patients (86 out of 529) had a combination of more than one test (**Table 2**). AJCH is a referral center for SMA rapid testing and gene therapy in the Middle East: 138 of our patients received rapid SMN1/2 analysis (**Supplementary Figure 1**), and a subset (N = 13) received additional testing.

**Table 2.** Testing Modality Per Clinical Indication

|  | NGS |  | CMA |  | P Value* | SMA |  | FragileX |  | MSMLPA/MLPA |  | Multiple Testing |  | Overall** |  |
| --- | --- | --- | --- | --- | --- | --- | --- | --- | --- | --- | --- | --- | --- | --- | --- |
|  | Total no. | %Positive (95% CI) | Total no. | %Positive (95% CI) |  | Total no. | %Positive (95% CI) | Total no. | %Positive (95% CI) | Total no. | %Positive (95% CI) | Total no. | %Positive (95% CI) | Total no. | %Positive (95% CI) |
| Clinical Indication |  |  |  |  |  |  |  |  |  |  |  |  |  |  |  |
| Neurological | 104 | 43.3%<br>(34.2 - 52.9) | 107 | 13.1%<br>(8.0 - 20.8) | 0.0001 | 138 | 34.8%<br>(27.3 - 43.0) | 44 | 2.3%<br>(0.4 - 11.8) | 10 | 70.0%<br>(39.7 - 89.2) | 75 | 33.3%<br>(23.7 - 44.6) | 320 | 34.7%<br>(29.7 - 40.1) |
| Non-Neurological | 163 | 39.9%<br>(32.7 - 47.5) | 53 | 24.5%<br>(14.9 - 37.6) | 0.048 | 0 | 0.0% | 0 | 0.0% | 4 | 25.0%<br>(4.6 - 69.9) | 11 | 36.4%<br>(15.2 - 64.6) | 209 | 37.3%<br>(31.0 - 44.1) |
| Overall | 267 | 41.2%<br>(35.5 - 47.2) | 160 | 16.9%<br>(11.9 - 23.4) | 0.0001 | 138 | 34.8%<br>(27.3 - 43.0) | 44 | 2.3%<br>(0.4 - 11.8) | 14 | 57.1%<br>(32.6 - 78.6) | 86 | 33.7%<br>(24.6 - 44.2) | 529 | 35.7%<br>(31.8 - 39.9) |

\* Comparisons between NGS versus CMA diagnostics yields. \*\* Overall count per patient, therefore, the sum/percentage will differ due to multiple tests performed per patient.

The analyses presented below focus on patients who received comprehensive genomic investigations (N = 427) for whom primary clinical indications were neurological/neurodevelopmental (30%), multisystem involvement (including nervous system) (28%), dysmorphic structural defects (10%) and inflammatory disorders (7.7%). The remaining 24% patients were referred for other primary indications including pulmonary, gastroenterology, sensory (vision and hearing), or haematological disorders (**Supplementary Table 3**).

### Genomic Testing Outcomes

Of the 529 probands, 189 received a positive molecular finding (**Supplementary Table 4**) for an overall diagnostic yield of 35.7% (95% CI, 31.7 – 39.9), which was higher for sequencing-based testing compared to chromosomal microarrays (41.2%; 95% CI, 35.5% - 47.2% versus 16.9%; 95% CI, 11.9% - 23.4%, respectively, P = 0.0001) across all indications (**Table 2**). Specifically, a molecular diagnosis was more likely to be obtained by sequencing relative to microarrays for patients presenting with neurological disorders (43.3%; 95% CI, 34.2% - 52.9% for sequencing versus 13.1%; 95% CI, 8.0% - 20.8% for microarrays, P = 0.0001) (**Supplementary Table 3**).

Of these 189 cases with positive findings, 29 (15.3%) were originally referred for a combination of different testing modalities, yet sequencing was required to identify diagnostic pathogenic variants in most of those cases (21 cases, 72.41%) (**Supplementary Table 4**).

Most patients with positive genetic findings were less than 5 years of age (71.9%). Younger patients between 0 and 2 years of age tended to have slightly higher diagnostic yield relative to patients who are 5 years or older (39.6%; 95% CI, 33.8% - 45.7% versus 30.8%; 95% CI, 24.4% - 38.1%, respectively, P = 0.066) (**Figure 1C** and **Table 3**).

**Table 3.** Molecular Diagnosis Rates by Age and

|  | Neurological |  | Non-Neurological |  | Overall* |  |
| --- | --- | --- | --- | --- | --- | --- |
|  | Total no. | %Positive (95% CI) | Total no. | %Positive (95% CI) | Total no. | %Positive (95% CI) |
| **Age, y** |  |  |  |  |  |  |
| 0 - 2 | 160 | 39.4%<br>(32.1 - 47.1) | 95 | 40.0%<br>(30.7 - 50.1) | 255 | 39.6%<br>(33.8 - 45.7) |
| 2 - 5 | 72 | 34.7%<br>(24.8 - 46.2) | 30 | 33.3%<br>(19.2 - 51.2) | 102 | 34.3%<br>(25.8 - 43.9) |
| ≥5 | 88 | 26.1%<br>(18.1 - 36.2) | 84 | 35.7%<br>(26.3 - 46.4) | 172 | 30.8%<br>(24.4 - 38.1) |
| **Overall** | 320 | 34.7%<br>(29.7 - 40.1) | 209 | 37.3%<br>(31.0 - 44.1) | 529 | 35.7%<br>(31.8 - 39.9) |

### Genetic Findings

124 rare diseases had an underlying genetic cause; the majority of these (N = 110) were observed only once. Thirteen disorders were seen 2 to 5 times (**Figure 1D** and **Supplementary Table 5**). All patients referred for SMA testing (N = 138), received rapid results within 4 days, on average, and a molecular diagnosis was made for 48 patients (35%) (**Figure 1D** and **Supplementary Figure 1**).

Of the 110 cases with positive findings by sequencing (**Supplementary Table 6**), 55 (50%) had biallelic pathogenic variants in genes associated with autosomal recessive disease (**Figure 1E**). Homozygous pathogenic variants were the most common in this group (46 out of 55 cases, 78.2%). On the other hand, 43 out of the 110 cases (39.1%) carried heterozygous pathogenic variants, including 6 confirmed de novo and 2 mosaic variants, in genes causing autosomal dominant disease. X-linked findings were reported in 7 cases (6.4%), and 5 patients (4.6%) had dual diagnoses involving a combination of autosomal recessive, autosomal dominant, and/or X-linked inheritance. Small exon-level copy number changes, detected by next generation sequencing read depth, were reported in 7 patients in this group (6.4%). A medically actionable secondary finding met ACMG criteria for reporting in one out of 79 families (1.27%) undergoing whole exome sequencing and consenting for receiving such findings (**Table 4** and **Supplementary table 6**). Finally, of the 125 clinically significant sequence variants identified in this group, 43 (34%) were novel or not previously reported in global disease databases (**Supplementary Table 6**).

**Table 4.** Molecular Findings

| Exome Sequencing |  |
| --- | --- |
| Mode of Inheritance | No. of Cases |
| Autosomal Dominant (n= 43) |  |
| Heterozygous SNV/INDELs |  |
| De novo | 6 |
| Inherited | 5 |
| Unknown | 29 |
| Mosaic Heterozygous SNV | 2 |
| Heterozygous CNV | 1 |
| Autosomal Recessive (n= 55) |  |
| Homozygous SNV/INDELs | 46 |
| Compound Heterozygous SNV/INDELs | 6 |
| Homozygous CNV | 3 |
| X-Linked Recessive/ X-Linked Dominant (n= 7) |  |
| Hemizygous CNV |  |
| Inherited | 1 |
| Unknown | 2 |
| Hemizygous SNV | 3 |
| Heterozygous SNV | 1 |
| Two Diagnosis (n= 5) |  |
| Autosomal Recessive + Autosomal Recessive | 1 |
| Autosomal Dominant + Autosomal Dominant | 1 |
| Autosomal Recessive + Autosomal Dominant | 1 |
| Autosomal Recessive + X-Linked | 1 |
| Autosomal Dominant + X-Linked | 1 |
| Secondary Diagnosis | 1 |
| Chromosomal Microarrays |  |
| Copy Number Variants (n= 24) |  |
| Heterozygous Deletion | 14 |
| Homozygous Duplication | 1 |
| Partial Trisomy | 1 |
| Partial Mosaic Tetrasomy | 1 |
| Whole Chromosome Trisomy | 5 |
| Whole Chromosome Monosomy | 1 |
| Suspected Unbalanced Translocation | 1 |
| Two Diagnosis (n= 1) |  |
| Heterozygous Deletion + Heterozygous Deletion | 1 |
| Loss of Heterozygosity (n= 2) |  |
| Uni Parental Disomy* | 2 |
| *UPD7 (case 212) and UPD15 (case 438) |  |

Of the 27 cases with positive microarray findings (**Supplementary Table 7**), 14 (51.9%) had single heterozygous pathogenic deletions. Chromosomal aneuploidies and uniparental disomies were detected in 6 and 2 cases, respectively. One patient had dual diagnoses due to two non-overlapping heterozygous deletions (**Table 4** and **Supplementary table 7**). Of the 160 cases referred for chromosomal microarrays, 59 (36.9%; 95% CI, 29.8% - 44.6%) had significant regions of homozygosity averaging around 7% of the autosomal genome (**Supplementary Table 8**), a finding consistent with the prevalent close relative marriages.

### Clinical Utility

Genetic findings offered new management and/or intervention plans for 124 out of the 189 cases (65.6%) with positive genetic findings across all ages (**Figure 1F** and **Supplementary Tables 6** and **7**). All 48 patients with diagnostic SMA findings were referred for gene therapy (ZOLGENSMA®). Excluding SMA, genetic findings uncovered new interventions and/or management plans for the remaining 76 out of 141 positive cases (53.9%) as summarized in **Supplementary Tables 6** and **7**. We also present case examples where genetic status positively altered treatment (**Appendix I**).

In addition to direct patient management, all families referred for genetic testing within AJCH (N = 425) were supported by certified genetic counsellors for test selection, pre-test counselling, reporting, and/or post-test counselling. Families were therefore informed about their recurrence risks and options for avoidance in future pregnancies.

### Impact on Diagnostic Odyssey

Although most patients with positive genetic findings were relatively young (**Figure 1C**), 35 of those patients (18.5%) were over 7 years of age (average 13 years, range 7 – 37 years) suggesting delayed diagnoses due to lack of access to genomic services in this region. We present a case example to highlight this issue.

Case 297 is a male in his 20’s with a long history of recurrent infections including eczema and ear infections during early childhood and recurrent pneumonias later in life leading to bronchiectasis. He had consistently elevated serum IgE and eosinophils throughout his course of illness and was symptomatically managed, including recurrent hospitalizations, without a clear working diagnosis. This patient was referred to AJCH for indication-based exome sequencing which revealed a homozygous pathogenic variant (c.4241+1G>A) in the DOCK8 gene which led to abnormal splicing (**Supplementary Figure 2**). This finding ended over a 10 year long diagnostic odyssey and patient was diagnosed with DOCK8-combined immunodeficiency syndrome, a life-threatening condition characterized by recurrent skin and respiratory infections, hyper IgE, and high risk of developing blood or skin cancers. No treatments were available for this condition.

Patient was then transferred to the National Institute of Health (NIH) where additional investigations revealed that he also has non-Hodgkin’s lymphoma. He then underwent allogenic hematopoietic stem cell transplant from a healthy sibling. Post-transplant, previously present eczematous dermatitis, recurrent infections, and lymphoma were all resolved.

## Discussion

We present a cohort of highly diverse patients with rare diseases from 41 countries primarily within the Middle East, Africa and Asia, a population which has been historically underserved in genomic services and underrepresented in global genetic studies. Our study demonstrates a high cumulative diagnostic yield (~36%) for genomic investigations in this cohort. This yield was highest for sequencing-based testing (~41%); recessive (autosomal and X-linked) inheritance accounted for ~60% of sequencing diagnoses in this study, consistent with the expected burden of single gene disorders in this population. Genomic sequencing testing was consequently the most effective testing strategy across several clinical indications. Genetic findings frequently led to early diagnoses, indicating new management and/or intervention plans, besides offering families information about recurrence risks and options to avoid disease in future pregnancies.

Around 37% of patients who received chromosomal microarray testing had significant regions of homozygosity spanning, on average, 7% of the autosomal genome. This finding is highly expected given the presence of close relative, including first cousin, marriages in the Middle East. Interestingly, 53 out of 133 patients (~40%) without conclusive findings by microarrays had such regions and are likely to be candidates for recessive novel gene discovery. In fact, we are currently pursuing further investigations of 8 cases with putative candidate genes underlying novel recessive disorders in this cohort (**Supplementary Table 9**).

Our overall sequencing-based testing diagnostic yield (41%) was similar to that reported in another study which focused on Saudi patients^7^, while our whole exome sequencing diagnostic yield was higher than that previously reported in other populations (34% versus 25% ^16,17^ – 29% ^18^). On the other hand, a substantial number of patients also received inconclusive results due to variants of uncertain clinical significance or VUSs (48.1%for WES and 32.6% for all sequencing tests) (**Supplementary Table 2**). This relatively high rate of VUS highlights the need for greater collection of sequencing data from this region – currently underrepresented in global genetic databases^5,6^ – which will allow us to better characterize both the ‘benign’ and ‘disease’ variation landscape, therefore improving variant classification and overall genetic data interpretation.

Despite the utility of genomic testing in our setting, this service is largely inaccessible to patients in the region due to lack of specialized centers with local genomic services. Furthermore, aside from Emirati citizens, who have free access to public healthcare services in the UAE, expatriate patients with private health insurance plans often do not get coverage for genetic testing. Of 373 genetic tests ordered for non-Emirati patients, the overall reimbursement rate was 16.4%, and was lowest for whole exome sequencing (8.2%). Limited understanding of the importance of genetic testing in the management of rare diseases in general, and specifically in this population as we show in this study, likely contributes to this low reimbursement rate, a trend we believe is prevalent in other countries with limited access to specialized services.

## Conclusions

Our study demonstrates that clinical genomic investigations should become standard of care for patients with rare diseases in this patient population. However, significant local investments are needed to establish multidisciplinary specialized centers where genomic investigations and subsequent management and intervention plans are accessible for patients with rare disorders. Furthermore, integrated research programs are essential to characterize novel disorders in this population and to expand the clinical annotation of the human genome. It is important to note that the reported diagnostic yield in this study is based on gene-disease associations primarily established in other populations. We therefore expect this yield will increase over time as more novel genes are characterized through study of this population. Establishing clinical genomics and research programs in this region will therefore not only benefit patients locally but will also enhance genomic data representation globally, expanding our understanding of the human genome.

## Data Availability

All data produced in the present work are contained in the manuscript

## Declarations

### Ethics approval and consent for publication

The de-identified, aggregate reporting in this study was approved by Dubai Healthcare Authority Research Ethics Committee (AJCH – 44).

### Availability of data and material

All data used and generated is available within the main manuscript and supporting files. Patients were not consented to share the raw data files beyond the research and clinical teams.

### Author’s Contribution

ME, SR, AND ANAT analysed the data. ME and ANAT drafted the manuscript. All authors edited the manuscript and approved the final version.

