## Supplementary figures and images for "The Genomic Landscape of Rare Disorders in the Middle East"

### Supplementary Figure 1

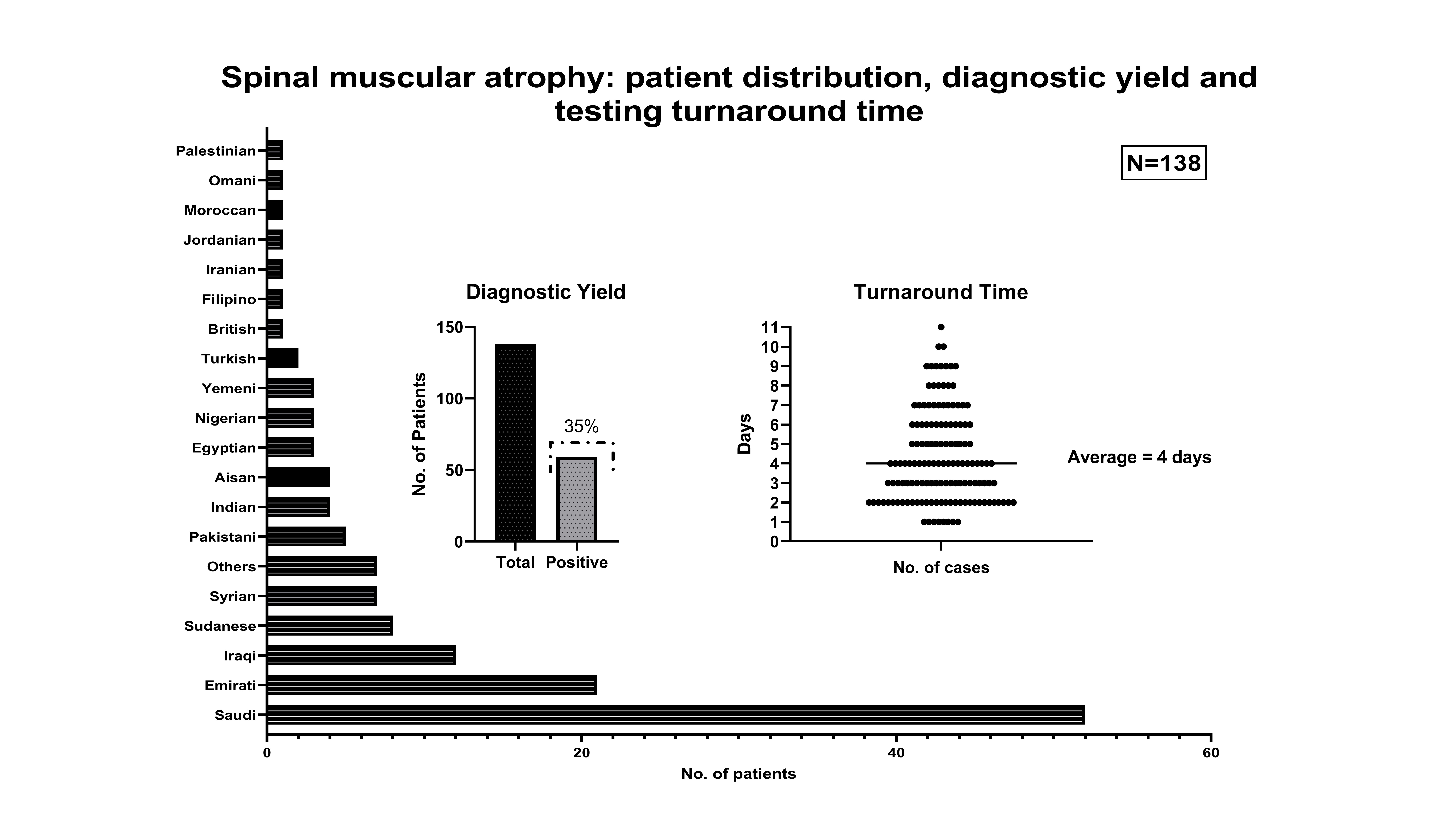

### Supplementary Figure 2

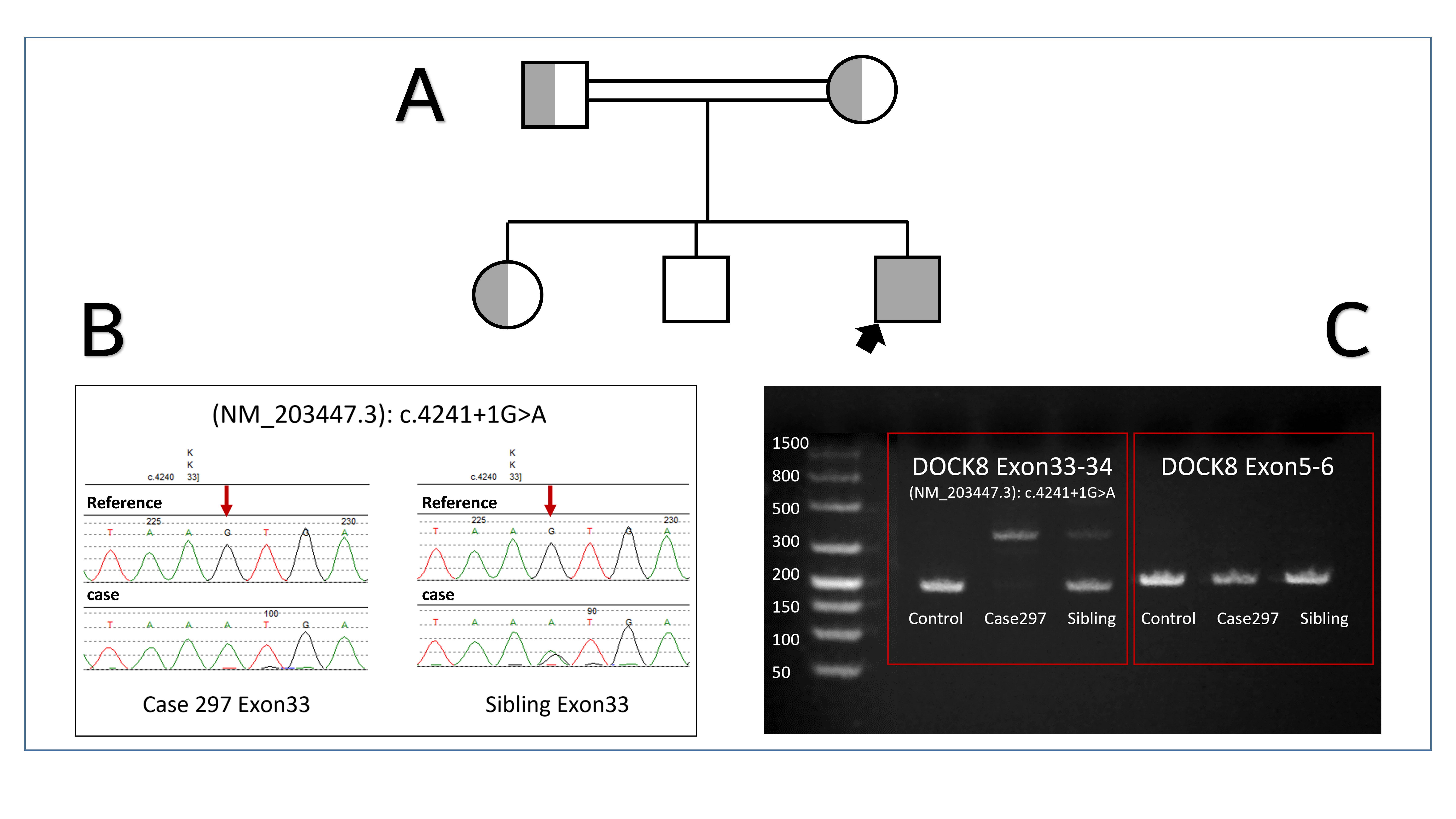
